# Chronic cluster headache: a study of the telencephalic and cerebellar cortical thickness

**DOI:** 10.1101/2021.03.01.21252083

**Authors:** Greta Demichelis, Chiara Pinardi, Luca Giani, Jean Paul Medina, Ruben Gianeri, Maria Grazia Bruzzone, Benjiamin Becker, Alberto Proietti, Massimo Leone, Luisa Chiapparini, Stefania Ferraro, Anna Nigri

## Abstract

Previous studies on brain morphological alterations in chronic cluster headache revealed inconsistent findings. The present cross-sectional explorative study determined telencephalic and cerebellar cortex thickness alterations in a relatively wide sample of chronic cluster headache patients (n=28) in relation to matched healthy individuals. The combination of two highly robust state-of-the-art approaches for thickness estimation (Freesurfer and CERES) with an unbiased functional characterization of the abnormal regions, revealed two main results. First, chronic cluster headache patients show cortical thinning in the right middle cingulate cortex and the left posterior insula. This indicates abnormalities in key-regions of pain processing areas, in particular in regions belonging to the spino-thalamic-cortical tract and primarily involved in the sensory-motor aspects of nociception. Second, chronic cluster headache patients present cortical thinning in the left anterior superior temporal sulcus and the left collateral/lingual sulcus, suggesting neuroplastic maladaptations in areas possibly involved in social cognition, which may promote psychiatric comorbidity, frequently observed in these patients.

## 1. Introduction

Cluster headache (CH) is a primary headache disorder characterized by episodes of short-lasting unilateral and severe craniofacial pain associated with trigeminal autonomic symptoms, such as lacrimation, ptosis, conjunctival injection, and facial sweating. About 90-95% of patients suffer from the episodic form of the disorder (eCH), characterized by periods of multiple daily episodes lasting weeks or months (in-bout phase - cluster period) followed by attack-free periods (out-of-bout phase - remission period). If the symptomatic period does not remit within 12 months, the disorder is considered chronic (cCH) (Headache Classification Committee of the International Headache Society, 2013).

Together with human hormonal and animal models studies (Leone and Bussone, 2009), neuroimaging promoted, in CH pathophysiology, a paradigmatic shift from peripheral pathogenetic hypotheses towards a central pathogenetic hypothesis, which emphasizes a possible key-role of the hypothalamus (May et al., 2018), and concomitant maladaptations in an additional range of pain-processing brain regions (Absinta et al., 2012; Rocca et al., 2010; Naegel et al., 2014; Giorgio et al., 2019). The seminal work of May et al. (May et al., 1999) described, for the first time, the anatomical rearrangement of the hypothalamus in CH patients. Although these results were not replicated in subsequent investigations (Matharu et al., 2006; Absinta et al., 2012; Naegel et al., 2014), a more recent study demonstrated an enlargement of the anterior portion of this structure in both eCH and cCH patients, suggesting a role in CH pathophysiology for the suprachiasmatic nucleus, the most important brain biological clock, and the paraventricular nucleus, a modulator of autonomic and nociceptive activities (Arkink et al., 2017).

Not fully consistent are also the results with respect to changes in grey matter (GM) volumes or thickness of pain processing regions, in particular across different CH phases (i.e., out-of-bout and in-bout) in eCH. During the out-of-bout phase, GM abnormalities were detected in the prefrontal and frontal areas (Absinta et al., 2012; Seifert et al., 2012; Giorgio et al., 2019), parietal lobe (Absinta et al., 2012; Seifert et al., 2012), temporal lobe, and insula cortex (Absinta et al., 2012; Naegel et al., 2014). Partially overlapping, GM alterations were observed during the in-bout phase specifically in frontal (Yang et al., 2013), insular (Naegel et al., 2014), and temporal (Naegel et al., 2014; Arkink et al., 2017) regions. Importantly, GM changes of extra pain-processing regions were also revealed, pointing to widespread neuromorphological changes in CH. In particular, the alterations within visual areas have been repeatedly reported in morphological (Absinta et al., 2012; Giorgio et al., 2019) and resting-state functional (rs-fMRI) (Rocca et al., 2010; Chou et al., 2017) studies.

Despite the importance of the comprehension of the neuropathological bases of CH chronicization, brain morphological investigations of the chronic condition are still very scarce and revealed conflicting results. On one side, Naegel et al. (2014) reported reduced GM volumes in core pain processing regions, such as the right (R) anterior cingulate cortex, the R somatosensory cortex, and the left (L) anterior insula cortex (IC), and in extra-pain processing regions, such as the L hippocampus, the L inferior temporal gyrus, the R orbitofrontal cortex, and the R occipital lobe. Moreover, these Authors observed increased GM volumes in the R posterior insula, the cerebellum, and the sensorimotor and visual areas suggesting, therefore, wide brain morphological rearrangements. On the other side, Arkink et al. (Arkink et al., 2017), in a large sample study, reported highly regional-specific increased GM volumes in the anterior section of the hypothalamus and the middle frontal gyrus, a region involved in the modulation of pain (Peyron et al., 2000).

Inconsistent findings between these studies may be explained in terms of the use of voxel-based morphometry (VBM) (Ashburner and Friston, 2000). One crucial theoretical assumption of VBM is that individual brain differences and anatomical correspondence of brain areas are maintained during the spatial normalization process (Pepe et al., 2014). However, this assumption is challenged by the spatial normalization inaccuracies (Bookstein, 2001; Senjem et al., 2005; Mechelli, 2005; Pepe et al., 2014) and the use of different spatial normalization templates and methods (Shen et al., 2007; Mechelli, 2005; Pepe et al., 2014), which strongly affect the results (Pepe et al., 2014). Moreover, VBM lacks a clear in-vivo or ex-vivo histological neurobiological validation in humans (Pepe et al., 2014). Differently, the Freesurfer’s (http://surfer.nmr.mgh.harvard.edu) (Dale et al., 1999) and the CERES’s automated pipelines (https://www.volbrain.upv.es) (Romero et al., 2017) are two robust, accurate, and widely accepted methods. The Freesurfer approach was validated using histological measures in autoptic brains (Rosas et al., 2002) and histological specimen in in-vivo brains (Cardinale et al., 2014), while the CERES was recently ranked first in the state-of-the-art methods for parcellating the cerebellum (Carass et al., 2018).

In an attempt to fill the gap among the reported inconsistencies and overcome the limitations of VBM, we investigated telencephalic and cerebellar cortex thickness abnormalities in a relatively wide sample of cCH patients compared to healthy individuals (CTRL) employing Freesurfer’s and CERES’s algorithms.

## 2. Materials and Methods

### 2.1. Participants

Twenty-eight cCH patients (23 men, 5 women; mean age ± SD: 45 ± 11.7 years; age range: 28-70 years; see table 1) were consecutively recruited among cCH individuals hospitalized to treat the recurrent CH attacks and assessed at the Fondazione IRCCS Istituto Neurologico C. Besta in Milan.

**Table 1:** Demographic and clinical data. Abbreviations: cCH = chronic cluster headache group, CTRL = control group, R = right, L = left, ys = years, n.a. = Not applicable/ not assessed.

|  |  | cCH | CTRL | Statistic | p-value |
| --- | --- | --- | --- | --- | --- |
| *Demographic data* | *N<sup>o</sup>* of participants | 28 | 28 |  |  |
| | Male/Female | 23/5 | 23/5 | $\chi^2(1, 56) = 7.66$ | p = 1.00 |
| | Age (M±SD) | 45 ± 11.7 (ys) | 45 ± 10.1 (ys) | $t(56) = 0.11$ | p = 0.74 |
| *Clinical data* | Side of attacks (R/L/bilat) | 15/12/1 | n.a. |  |  |
| | *N<sup>o</sup>* of daily attacks (M±SD) | 5.0 ± 2.4 | n.a. | | |
| | Duration of cCH (M±SD) | 6.9 ± 6.3 (ys) | n.a. | | |
| *Daily acute treatment* | Sumatriptan posology (M±SD) | 4 ± 2.2 (tablet, 100mg) | n.a. | | |
| *Current profilaxys* | Anticonvulsants (Valproate, Topiramate) | 7 (25%) | n.a. |  |  |
|  | Lithium | 13 (46.%) | n.a. |  |  |
|  | Calcium-channel blockers (Verapamil) | 16 (57.1%) | n.a. |  |  |
|  | Antidepressants (Duloxetine, Amitriptyline) | 1 (3.6%) | n.a. |  |  |
|  | Corticosteroids (Dexamethasone) | 7 (25%) | n.a. |  |  |
|  | Other | 1 (3.6%) | n.a. |  |  |
| *Tobacco use* | *N<sup>o</sup>* of tobacco smokers (>1/day) | 23 (23 Male/0 Female) | n.a. |  |  |
| | *N<sup>o</sup>* cigarettes/day | 18 ± 10.6 (M±SD) | n.a. | | |
| | *N<sup>o</sup>* of heavy tobacco smokers ($\geq 25/day$) | 7/23(30%) | n.a. | | |

All patients were diagnosed with cCH by two expert neurologists (M.L. and A.P.) according to the International Classification of Headache Disorders, 3rd edition criteria (ICHD-3) (Headache Classification Committee of the International Headache Society, 2013). Patients with a concomitant diagnosis of other primary or secondary headache disorders, cardiovascular diseases, diabetes mellitus, hypertension, or a history of psychiatric conditions were excluded from the study.

Except for two, all patients were taking prophylactic treatment (table 2).

**Table 2:**
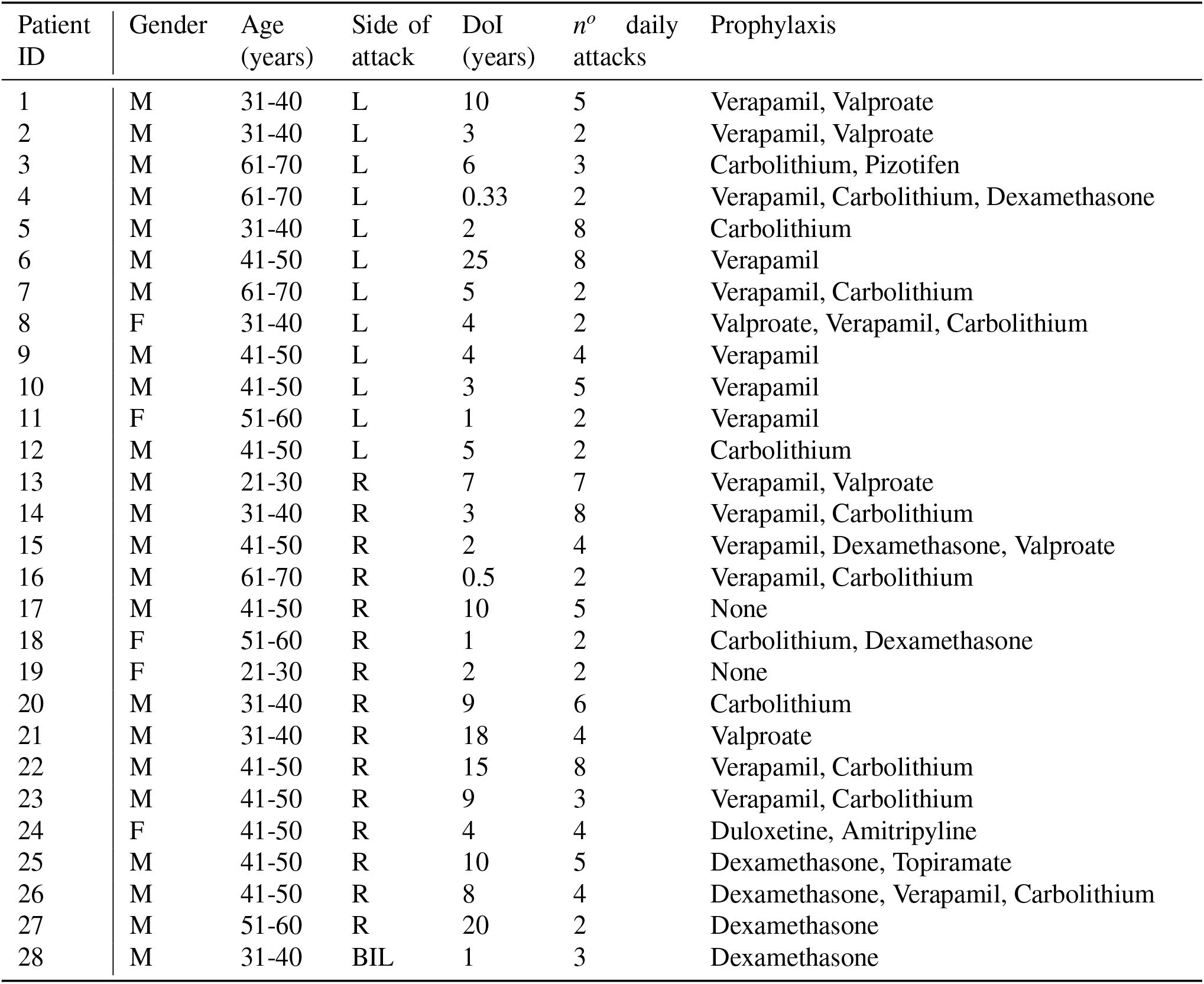
Current prophylactic treatment in cCH patient. Abbreviations: M = male, F= female, L = left, R = right, BIL = bilateral, DoI = duration of illness, ys = years.

Twenty-eight sex and age-matched healthy individuals (CTRL) (23 men, 5 women; mean age ± SD: 45 ± 10.1 years; age range: 29-64 years), reporting no history of primary headache or chronic pain and no history of neurological or psychiatric disorders, were recruited as controls.

All participants provided written informed consent according to the Declaration of Helsinki before the study inclusion. This study was approved by the Fondazione IRCCS Istituto Neurologico C. Besta ethics committee.

### 2.2. MRI assessments

All participants were scanned with a 3T magnetic resonance imaging (MRI) system (Achieva, Philips Healthcare BV, Best, NL) using a 32-channel head coil. As part of the MRI protocol, high-resolution structural 3D T1-weighted (T1w) image (TR = 9.86 ms, TE = 4.59 ms, FOV = 240×240 mm, voxel size = 1 mm^3^, flip angle = 8^°^, 185 sagittal slices) was acquired in each participant. All MRI images were visually inspected by expert neuroradiologists (L.C. and M.G.B.) to exclude apparent brain abnormalities or artifacts affecting the quality of the exam.

### 2.3. Structural MRI data analyses

We examined the presence of cerebral structural abnormalities in cCH patients through investigating the cortical thickness of telencephalic regions and cerebellar lobes, using automated, well-validated, and robust tools (respectively, Freesurfer and CERES) (Rosas et al., 2002; Fischl, 2012; Cardinale et al., 2014; Romero et al., 2017).

#### Telencephalic cortex thickness analyses

We performed surface-based analyses, employing Freesurfer version 6 (Massachusetts General Hospital, Harvard Medical School; http://surfer.nmr.mgh.harvard.edu). In particular, cortical model reconstructions were performed using the *recon-all* pipeline, an automated algorithm. Briefly, for each subject, after the normalization to the MNI305 space and intensity signal normalization, automated cortical segmentation was applied on the T1w image. The segmentation output was visually checked and manually corrected by an expert operator (C.P.) blind to the diagnosis (cCH or CTRL) to remove the pial-white boundary surfaces segmentation inaccuracies. A surface was then created covering the hemispheres with triangles (i.e. tesselation), and applying a topology fixer and a spatial smoothing (10mm FWHM). The obtained individual cortical models were registered with a folding-patterns spherical atlas (Fischl et al., 1999), to match the individual patterns of cortical folding across participants.

The resulting surfaces were then subjected to a vertex-wise between-group analysis to test for cortical thickness differences between cCH and CTRL individuals. The estimated total intracranial volumes (eTIV), obtained as Freesurfer output files, were used as covariates in the analysis. The results were considered significant with a vertex-wise threshold of p<0.001 and a cluster-wise threshold of 50 mm^2^. Significant clusters were anatomically labeled according to the Destrieux Cortical Atlas (Destrieux et al., 2010). Coordinates of the peaks of the observed cortical abnormalities were transformed from MNI305 to MNI152 using the matrix provided in https://surfer.nmr.mgh.harvard.edu/fswiki/CoordinateSystems.

#### Cerebellar cortex thickness analyses

We computed the cerebellar cortical thickness using the automated pipeline CERES software (https://www.volbrain.upv.es) (Romero et al., 2017). Briefly, each subject’s T1w image was denoised, corrected for intensity inhomogeneities, and registered to the MNI152 space. The image was then cropped and normalized to the MNI152 cerebellum atlas. After the preprocessing, CERES applied an automated cerebellum patch-based segmentation and produced as output the mean cortical thickness of each cerebellar lobule. To reduce the number of multiple comparisons when performing the subsequent analyses (see beyond), we computed the mean cerebellar cortical thickness of the bilateral anterior (I, II, III, IV, and V lobules), posterior (VI, VII, VIII, IX lobules, and crus I and II) and flocculonodular (X lobule) lobes (D’Mello and Stoodley, 2015).

To identify possible cerebellar cortex differences, we compared the mean cerebellar cortex thickness of each lobe between cCH and CTRL individuals. Shapiro-Wilk test showed that the data were normally distributed and Barrett test that the variance between groups was no statistically different, therefore, two-tailed t-tests, were employed.

For all the above tests, the results were considered significant with a p<0.05 corrected with the Holm-Bonferroni method (Holm, 1979).

#### Functional characterization of the abnormal cortical regions

To provide an unbiased functional characterization of the clusters of cortical abnormalities observed in our sample of cCH patients, we employed NiMARE (https://nimare.readthedocs.io/en/latest/), and Neurosynth, an online platform to synthesize large-scale functional magnetic resonance imaging (fMRI) data (http://neurosynth.org) (Yarkoni et al., 2011), which presents, to date, 14371 fMRI studies for a total of 507891 activations.

First, we explored the possibility that the clusters of cortical abnormalities were functionally connected. Employing the Neurosynth function *‘locations’*, we generated resting-state functional connectivity (rs-FC) maps (thresholded at r>0.2) seeded in each peak coordinates of the observed clusters of cortical abnormality. These rs-FC maps were obtained from the rs-fMRI data analysis of 1000 human subjects (provided in Neurosynth as a courtesy of T. Yeo, R. Buckner, and the Brain Genomics Superstructure Project) (Buckner et al., 2011; Yeo et al., 2011). Then, we determined whether each single rs-FC map comprised, beyond the seed used to generate it, the other clusters of cortical abnormalities. We built a 10mm spherical region of interest (ROI) around the peak coordinates of each cluster of abnormality and we extracted the percentage of the ROI voxels overlapping each rs-FC map. When at least 50% of the ROI voxels overlapped the rs-FC map, we considered that cluster of cortical abnormalities (from which the ROI was built) as functionally connected to the cluster used as seed to build that rs-FC map.

Second, based on the observed regions of cortical thinning, we identified the possible cognitive processes that might be impaired in CH pathophysiology. Using NiMARE (function *‘nimare.decoder.discrete’*), we identified which terms were specifically employed in studies present in the Neurosynth database, that reported fMRI activity in the previous generated ROIs. This approach allowed us to define whether activation in the investigated ROIs was preferentially related to specific terms, a proxy of cognitive processes (Yarkoni et al., 2011).

From the output of the decoder, we excluded terms reporting anatomical regions and generic terms (such as ‘networks involved’, ‘nervous’, ‘spatial temporal’) and we arbitrarily decided to consider the first 20 terms ordered in descending order according to the conditional probability of a term being used in a study given that the activation is present in that ROI [*P*(*term* | *activation*)] (i.e. posterior probability). We then identified similar terms, possibly indicating common cognitive processes, related to the clusters defined as functionally connected based on the previous analysis.

## 3. Results

### Telencephalic cortex thickness results

The between-group vertex-wise analysis showed a reduced cortical thickness in cCH patients compared to CTRL individuals, located in four distinct areas: the R midcingulate cortex (MCC), mainly in its posterior part; the L superior temporal sulcus (STS), mainly in its anterior part; the L collateral sulcus/lingual sulcus (CLS); and the L posterior insula cortex (post-IC) (see figure 1and table 3).

**Table 3:** Vertex-wise between-group analysis: significant clusters of cortical thickness reduction in cCH patients compared to CTRL (p <0.001, cluster-wise threshold = 50 *mm*^2^). Significant regions were labelled according to the Destrieux Cortical Atlas (Destrieux et al., 2010). Peak-coordinates in MNI152 space. Abbreviations: ant. = anterior, post. = posterior, R = right, L = left.

| Region | peak-coordinates | | | $z$-score | Area ($\text{mm}^2$) | $p$-value |
| --- | --- | --- | --- | --- | --- | --- |
|  | x | y | z |  |  |  |
| R midcingulate cortex | +14 | +3 | +45 | -4.68 | 59.6 | <0.0001 |
| L superior temporal sulcus | -50 | -6 | -21 | -4.79 | 152.4 | <0.0001 |
| L collateral/lingual sulcus | -32 | -40 | -6 | -3.73 | 69.8 | 0.0001 |
| L posterior insula cortex | -33 | -33 | +18 | -3.47 | 76.5 | 0.0003 |

### Cerebellar cortex thickness results

The two-tailed t-tests showed no significant differences between cCH patients and CTRL individuals in cerebellar cortex thickness (see table 4).

**Table 4:** Mean and standard deviation of cerebellar lobe cortical thickness (in mm) and results from the between-group comparison (cCH vs. CTRL individuals) using two-tailed t-tests (p<0.05 after Holm-Bonferroni correction). Abbreviations: cCH = chronic cluster headache patients, CTRL = control individuals.

| Region | cCH | CTRL | p-value |
| --- | --- | --- | --- |
| Anterior Lobe | $3.73 \pm 0.19$ | $3.82 \pm 0.14$ | 0.11 |
| Posterior Lobe | $4.66 \pm 0.17$ | $4.72 \pm 0.20$ | 0.41 |
| Flocculonodular Lobe | $2.94 \pm 0.27$ | $2.89 \pm 0.26$ | 0.49 |

### Functional characterization of the abnormal cortical regions

The peak coordinates of the 4 clusters of cortical thinning that emerged in CH patients (i.e., R MCC, L postIC, L STS, and L CLS) were used to build the respective rs-FC maps and the 10mm spherical ROIs. The extraction of the percentages of the voxels of the ROIs overlapping the rs-FC maps of interest showed that 92.2% of the ROI voxels in the L postIC overlapped the rs-FC map seeded in the R MCC and, conversely, that 78.1% of the ROI voxels in the R MCC overlapped the rs-FC map seeded in L postIC. Similarly, 81.6% of the L STS ROI voxels overlapped the rs-FC map seeded in the L CLS cluster, and 78.5% of the L CLS ROI voxels overlapped the L STS rs-FC map (table 5). Based on these results, we inferred that functional connectivity occurred between the R MCC and the L postIC and between the L STS and the L CLS.

**Table 5:** Percentage of the voxels in the regions of interest (ROIs) overlapping the resting-state functional connectivity maps seeded in right midcingulate cortex (R MCC), left posterior insula cortex (L postIC), left superior temporal sulcus (L STS) and left collateral/lingual sulcus (L CLS). Resting-state functional connectivity maps were built in Neurosynth, employing the resting-state fMRI data collected in 1000 individuals, as a courtesy of T. Yeo, R. Buckner, and the Brain Genomics Superstructure Project. (Yeo et al., 2011; Buckner et al., 2011)

| ROI coordinates (x,y,z) | Network rs-FC seeded in |  |  |  |
| --- | --- | --- | --- | --- |
|  | R MCC | L postIC | L STS | L CLS |
| 14 4 44 ( R MCC) | 100% | 78.1% | 4.45% | 15.9% |
| -32 -32 18 (L postIC) | 92.2% | 100% | 38.6% | 44.7% |
| -50 -6 -20 (L STS) | 13.2% | 48.9% | 100% | 81.6% |
| -32 -40 -6 (L CLS) | 2.2% | 29% | 78.5% | 100% |

Capitalizing on these evidences, we observed that the common terms between the R MCC ROI and the L postIC ROI were related to pain processing, while the common terms between L STS and L CLS ROI were mainly related to the so-called ‘social brain’ (Spreng and Mar, 2012; Frith and Frith, 2012). 6.

Specifically, the output of the decoder applied in the R MCC ROI (table 6) showed a posterior probability of 0.69 for the terms ‘noxious’ and ‘nociceptive’, thus indicating that 69% of the studies in the Neurosynth database including these terms presented fMRI activity in the cluster of cortical thinning of the R MCC. The output of the decoder applied in the L postIC ROI (table 6) presented one term referred to pain processing (‘noxious’) with a posterior probability of 0.73, therefore indicating that 73% of the studies including this term presented fMRI activity in this cluster of cortical thinning (table 6).

**Table 6:** Decoder results obtained using the NiMARE function *‘nimare.decoder.discrete’* in 10mm spherical regions of interest (ROIs), centered in the peaks coordinates of the clusters of cortical thinning (right midcingulate cortex and left posterior insula cortex) observed in cluster headache patients. Abbreviations: ROI = region of interest, R MCC = right midcingulate cortex, L postIC = left posterior insula cortex, pReverse = p-value of reverse inference. zReverse = z value of reverse inference, probReverse = posterior probability, * common terms for the two ROIs.

| Terms | Reverse metrics |  |  |
| --- | --- | --- | --- |
| ROI in R MCC | pReverse | zReverse | probReverse |
| motor imagery | 0.0000 | 6.12 | 0.77 |
| extension | 0.0001 | 4.03 | 0.73 |
| tapping | 0.0000 | 4.82 | 0.73 |
| finger tapping | 0.0003 | 3.66 | 0.73 |
| focusing | 0.0000 | 4.16 | 0.71 |
| *nociceptive | 0.0054 | 2.78 | 0.70 |
| *noxious | 0.0028 | 2.99 | 0.69 |
| executive functions | 0.0005 | 3.49 | 0.69 |
| motor control | 0.0000 | 4.40 | 0.69 |
| practice | 0.0000 | 4.25 | 0.69 |
| individually | 0.0030 | 2.96 | 0.69 |
| execution | 0.0000 | 5.66 | 0.69 |
| externally | 0.0039 | 2.88 | 0.68 |
| saccade | 0.0123 | 2.50 | 0.68 |
| constructed | 0.0123 | 2.50 | 0.68 |
| muscle | 0.0070 | 2.70 | 0.68 |
| tones | 0.0070 | 2.70 | 0.68 |
| monitor | 0.0079 | 2.66 | 0.68 |
| finger | 0.0000 | 4.75 | 0.67 |
| monitor | 0.0079 | 2.66 | 0.68 |
| ROI in L postIC |  |  |  |
| pitch | 0.0000 | 6.33 | 0.81 |
| multisensory | 0.0000 | 7.19 | 0.80 |
| sounds | 0.0000 | 8.21 | 0.80 |
| speech production | 0.0000 | 5.58 | 0.79 |
| vocal | 0.0000 | 4.91 | 0.79 |
| spectral | 0.0000 | 4.82 | 0.78 |
| musical | 0.0000 | 5.77 | 0.78 |
| hearing | 0.0000 | 5.35 | 0.78 |
| sound | 0.0000 | 7.64 | 0.78 |
| speech perception | 0.0000 | 4.45 | 0.77 |
| audiovisual | 0.0000 | 4.63 | 0.76 |
| speaker | 0.0003 | 3.60 | 0.76 |
| pressure | 0.0001 | 3.96 | 0.75 |
| acoustic | 0.0000 | 5.06 | 0.75 |
| listening | 0.0000 | 5.87 | 0.75 |
| auditory | 0.0000 | 8.21 | 0.75 |
| music | 0.0000 | 4.48 | 0.74 |
| silent | 0.0011 | 3.26 | 0.74 |
| *noxious | 0.0017 | 3.14 | 0.73 |
| paced | 0.0017 | 3.13 | 0.72 |

The output of the decoder applied in the L STS ROI (table 7) presented 7 terms possibly referred to ‘social-brain’ processes (‘mind tom’, ‘mentalizing’, ‘tom’, ‘mental states’, ‘theory of mind’, ‘autobiographical’, ‘autobiographical memory’) (Spreng and Mar, 2012; Frith and Frith, 2012) with high posterior probabilities (0.75-0.79), while the output of the decoder applied in the L CLS ROI (table 7) presented mainly terms related to memory process (e.g. ‘remembered’, ‘recollection’, ‘construction’), but also terms with high posterior probability (0.77) related to the ‘social brain’, such as ‘autobiographical’ and ‘autobiographical memory’ (table 7).

**Table 7:** Decoder results obtained using the NiMARE function *‘nimare.decoder.discrete’* in 10mm spherical regions of interest (ROIs), centered in the peaks coordinates of the clusters of cortical thinning (left superior temporal sulcus and left collateral/lingual sulcus) observed in cluster headache patients. Abbreviations: ROI = region of interest, R STS = left superior temporal sulcus, L CLS = left collateral/lingual sulcus, pReverse = p-value of reverse inference. zReverse = z value of reverse inference, probReverse = posterior probability, common terms for the two ROIs.

| Terms | Reverse metrics |  |  |
| --- | --- | --- | --- |
| ROI in L STS | pReverse | zReverse | probReverse |
| *construction | 0.0000 | 6.83 | 0.80 |
| *autobiographical | 0.0000 | 8.21 | 0.79 |
| mind tom | 0.0000 | 5.85 | 0.79 |
| *autobiographical memory | 0.0000 | 6.16 | 0.79 |
| tom | 0.0000 | 5.46 | 0.78 |
| language comprehension | 0.0000 | 6.16 | 0.78 |
| comprehension | 0.0000 | 8.21 | 0.77 |
| sentences | 0.0000 | 8.21 | 0.77 |
| mentalizing | 0.0000 | 6.75 | 0.77 |
| sentence | 0.0000 | 8.21 | 0.76 |
| mental states | 0.0000 | 6.11 | 0.76 |
| *remembering | 0.0000 | 4.39 | 0.75 |
| language network | 0.0000 | 4.34 | 0.75 |
| theory mind | 0.0000 | 6.26 | 0.75 |
| negative neutral | 0.0000 | 4.29 | 0.74 |
| speaker | 0.0000 | 4.13 | 0.74 |
| syntactic | 0.0000 | 5.71 | 0.74 |
| verbs | 0.0000 | 4.50 | 0.74 |
| heard | 0.0002 | 3.75 | 0.73 |
| sentence comprehension | 0.0002 | 3.69 | 0.73 |
| ROI in L CLS |  |  |  |
| navigation | 0.0000 | 6.92 | 0.81 |
| *remembered | 0.0000 | 8.21 | 0.79 |
| recollection | 0.0000 | 7.63 | 0.78 |
| mental imagery | 0.0000 | 5.51 | 0.77 |
| *autobiographical memory | 0.0000 | 5.51 | 0.77 |
| *autobiographical | 0.0000 | 6.87 | 0.77 |
| virtual | 0.0000 | 7.17 | 0.77 |
| scenes | 0.0000 | 8.21 | 0.77 |
| *remembering | 0.0000 | 4.70 | 0.76 |
| episodic memory | 0.0000 | 8.21 | 0.75 |
| retention | 0.0000 | 4.46 | 0.75 |
| episodic | 0.0000 | 8.21 | 0.75 |
| scene | 0.0000 | 6.28 | 0.75 |
| encoding retrieval | 0.0000 | 5.15 | 0.74 |
| young older | 0.0001 | 3.82 | 0.74 |
| place | 0.0000 | 5.76 | 0.73 |
| *construction | 0.0001 | 3.82 | 0.73 |
| details | 0.0000 | 4.18 | 0.73 |
| memory encoding | 0.0000 | 4.99 | 0.73 |
| recognition memory | 0.0000 | 4.91 | 0.72 |

## 4. Discussion

The present study capitalized on two robust and extensively validated methods (Freesurfer and CERES pipelines applied on T1-3D MR images) to determine telencephalic and cerebellar cortex structural abnormalities in the chronic form of CH. Our study revealed that cCH patients compared to CTRL individuals presented cortical thinning in regions belonging to two functionally separable brain systems: the R MCC and in the L post-IC, regions playing a key role in pain processing (Peyron et al., 2000), and the L STS and L CLS, extra-pain processing areas, possibly part of the ‘social brain’ (Frith and Frith, 2012).

To strengthen our inferences, we functionally characterized these cortical areas using an unbiased large-scale dataset approach (Yarkoni et al., 2011), which supports the hypothesis that these clusters of cortical thinning belong to two different functional systems mainly involved in pain-related and, possibly, social-related processes.

### 4.1. Cortical thinning of R MCC and L postIC

In cCH patients, Naegel et al. (2014) reported reduced volume of the R posterior-anterior cingulate cortex (in a region close but non-overlapping to the one we observed), and increased volume in the R postIC.Conversely, Arkink et al. (Arkink et al., 2017) did not detect any sign of abnormalities in the IC or cingulate cortex in cCH patients. However, alterations of the insula cortex and cingulate cortex have been previously observed in eCH patients but in different areas compared to our findings (Absinta et al., 2012; Yang et al., 2013).

It is well accepted that R MCC and L postIC play an important role in the experience of pain (Legrain et al., 2011; Wager et al., 2013; Garcia-Larrea and Peyron, 2013). These are core-areas of the spino-thalamic-cortical tract, one of the two main ascending systems (the other being the spino-parabrachial-amygdalar pathway) that convey and process nociceptive information. This pathway, after the thalamic relay, projects to the postIC, the MCC, parietal regions, and, to a lesser extent, to primary sensory and motor areas (Dum et al., 2009; Lenz et al., 2010). Accordingly, the identified clusters of cortical thinning in the R MCC and the L postIC appeared to be functionally connected, as reflected by the seed-based analyses in a large independent rs-fMRI dataset (Yeo et al., 2011; Buckner et al., 2011). This is also confirmed by previous investigations showing that the posterior MCC is functionally connected to the postIC (Cauda et al., 2011; Taylor et al., 2009). Moreover, our results from the application of the decoder across the whole Neuroynth database indicated that studies showing activity in these areas present high posterior probability of presenting terms referred to pain processing.

The MCC has been implicated in a wide range of cognitive functions associated with nociception, including monitoring, cognitive conflict, reward, and emotional and aversive stimuli processing (Vogt, 2016; Heilbronner and Hayden, 2016; Shackman et al., 2011; Etkin et al., 2011; Kober et al., 2008). This region is subdivided into an anterior and a posterior section (Vogt, 2016). The anterior MCC was consistently shown to be involved in the processing of the negative affects during anticipation or experience of pain and aversive stimuli (Vogt, 2005), while the posterior MCC (where we mainly detected the cortical thinning - see figure 1), is involved in the reflexive spatial orientation of the head and body towards salient sensory stimuli (Vogt, 2009) acting as an interface between cognition and action (Heilbronner and Hayden, 2016).

**Figure. 1:**
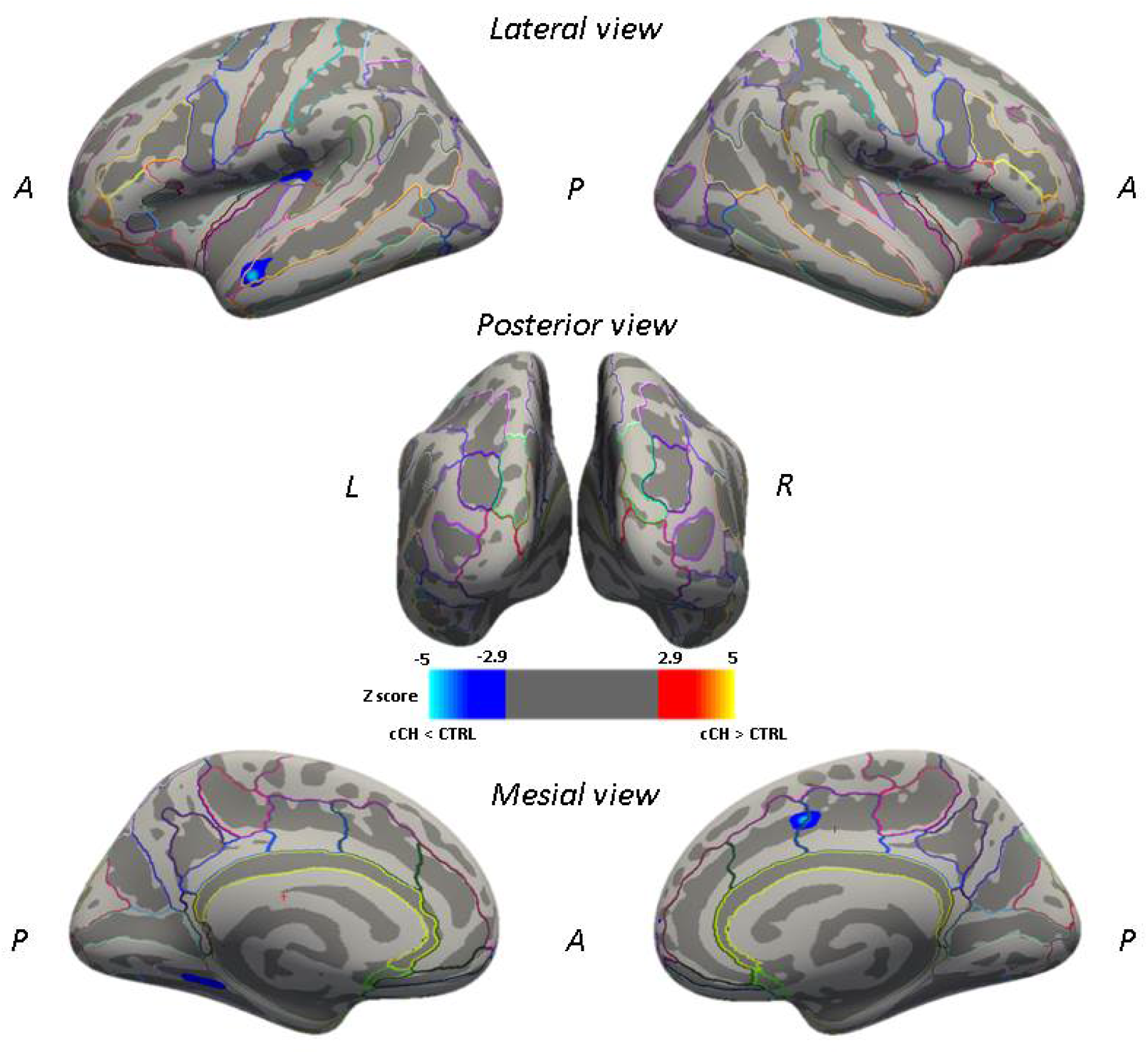
Vertex-wise between-group analysis: significant clusters of reduced cortical thickness in cCH patients compared to CTRL participants expressed as z-values (p <0.001, cluster-wise threshold 50 *mm*^2^), displayed on FreeSurfer’s semi-inflated cortical surfaces. The color bar indicates the significance levels of the clusters. Abbreviations: cCH = chronic cluster headache patients, CTRL = control individuals.

The postIC codes the intensity and the location of the nociceptive inputs and in both human (Frot et al., 2007; Baumgärtner et al., 2010; Henderson et al., 2007; Mazzola et al., 2009; Segerdahl et al., 2015) and non-human primates (Craig, 2014) its main role is to represent the sensory features of the nociceptive inputs. This key-role in the sensory processing of nociception is also underlined by acute painful sensations induced by its stimulation (Ostrowsky et al., 2002) and by the alteration of pain processing when this region is lesioned (Garcia-Larrea and Peyron, 2013; Birklein et al., 2005). From this perspective, our results indicate abnormalities in areas mainly involved in the sensory-motor processing of nociceptive information traveling along the spino-thalamic pathway.

In a recent murine study the development and maintenance of nociceptive hypersensitivity, important in chronic pain, is mediated by the MCC-post-IC pathway (Tan et al., 2017): the inactivation of the MCC blocked the development and maintenance of the secondary hypersensitivity, characterized by a relevant ongoing activity of the MCC but did not alter the acute pain perception. Moreover, the inactivation of the MCC projections to the postIC, and from the postIC to the raphe magnus, where descending modulatory systems are located, aborted the development of the secondary hypersensitivity. These results indicate that the sustained activity of the MCC induces plastic changes promoting and maintaining hypersensitivity and cortical sensitization, also modulating the postIC relay and the descending modulatory systems to the raphe nuclei. It is tempting to speculate that the reduced cortical thickness of MCC and postIC we observed in cCH patients might indicate a dysfunctional adaptation of this network that can be critical also in CH chronicization.

### 4.2. Cortical thinning of L STS and L CLS

Converging evidence has shown that STS is a fundamental hub for social cognition, being involved in the perception of faces (Puce et al., 1996), voices (Belin et al., 2000), biological motions (Allison et al., 2000; Pelphrey et al., 2005), and in the inference of others’ mental states and intention of others’ actions (Vander Wyk et al., 2009; Pelphrey et al., 2004; Ciaramidaro et al., 2007). It has been proposed that different STS subregions host diverse social cognition processes.

In particular, the anterior section of STS, detected as thinned in our sample of cCH patients (figure1), is part of the so-called ‘social brain’ (Burnett and Blakemore, 2009; Simmons et al., 2010; Pascual et al., 2015), a wide network comprising also the lateral fusiform gyrus, the medial prefrontal cortex, the precuneus, the posterior STS, and the amygdala (Mellem et al., 2016). The STS area was consistently shown to be involved in the representation of the social/emotional concepts (Wang et al., 2019) and the processing of the emotional state of faces (Schobert et al., 2018). Supporting this notion, the results from the application of the decoder across the whole Neuroynth database, revealed that studies presenting fmri activity in this area show very high probability of presenting terms referred to ‘social brain’ and, in particular, to autobiographical memories processes. Despite specific GM reductions of the L anterior STS were not reported in previous studies on CH, abnormalities in the inferior temporal lobe in cCH patients (Naegel et al., 2014) and the middle temporal gyrus in eCH patients (Arkink et al., 2017) were reported.

We have also found evidence of reduced cortical thickness in the L CLS. Importantly, this region appears to be functionally connected with L STS, as emerged from the seed-based analyses in a large independent rs-fMRI dataset (Yeo et al., 2011). Notably, also this area can be considered part of the ‘social brain’: according to the results of the decoder, when a fMRI work reports this area as active, terms referring to the ‘social brain’ (i.e. ‘autobiographical’ and ‘autobiographical memories’) show high probability to be present.

We speculate that the thinned regions of the anterior section of the L STS and the L CLS, observed in our sample of cCH patients, might belong to the ‘social brain’ network This network was reported to be dysfunctional in several neuropsychiatric conditions, such as major depression, schizophrenia, and Alzheimer’s disease (Porcelli et al., 2019). Depression, anxiety and aggressive behaviour are the most common psychiatric comorbidities of CH (Robbins, 2013; Louter et al., 2016), therefore, a vulnerability of the ‘social brain’ in CH pathophysiology seems plausible.

### 4.3. Limitations

Two main limitations suggest caution in the interpretation of these data. The first is related to the ongoing pharmacological treatment of the investigated cCH patients that might impact the brain structure. For example, chronic corticosteroid therapy can reduce the volumes of the hippocampus and the amygdala (Dieleman et al., 2017). For ethical reasons, we did not ask the patients to discontinue their medications to perform the washout.

The second limitation is related to the fact that we did not match the cCH and CTRL individuals for smoking habits. Heavy smoking habits are consistently reported in CH. Although the relationship between the two is still unrevealed, it was hypothesized a causal role of smoke in the development of CH (Manzoni, 1998; Rozen, 2010) or that specific personality traits (Lambru et al., 2010) make these individuals more vulnerable to certain unhealthy habits and/or substance abuse/addiction. It is well acknowledged that smoking-dependent individuals present morphological abnormalities of several brain regions (Yang et al., 2020). Overlapping with our results, a recent metaanalysis showed that smokers, in comparison to non-smokers, present reduced grey matter volumes in the L postIC (Yang et al., 2020). This observation underlines the need to consider this variable in future studies to understand the interactions between smoke and CH pathophysiology in producing brain alterations.

### 4.4. Conclusion

In this study, two robust automatic tools at the state of the art (Freesurfer and CERES) have been used to evaluate the telencephalic and cerebellar cortical thickness in a relatively wide sample of cCH patients.

We show that these patients present cortical thinning in brain regions belonging to the spino-thalamic-cortical tract and mainly involved in the sensory-motor processing of pain (R MCC and L postIC), possibly supporting the chronicization of CH. Moreover, we show that cCH patients present reduced cortical thickness in areas belonging to the ‘social brain’ (L anterior STS and posterior CLS), and this could be in relation with the psychiatric comorbidity frequently observed in cCH individuals (Robbins, 2013). Future studies should assess whether, in cCH individuals, the ‘social brain’ is impacted from the behavioral point of view, employing neuropsychological tests investigating, for example, the integrity of the theory of mind or of emotion recognition, and from the neural point of view, employing specific fMRI cognitive tasks. Despite these important results, we could not ascertain if these abnormalities are the cause or the effect of cCH. However, our study paves the way for hypothesis-driven studies, that might impact our understanding of cCH pathophysiology.

## Data Availability

Data are available upon request to the corresponding author.

## Funding sources

This work was supported by the Italian Ministry of Health, research grant RF-2016-02364909.

## Conflict of interest

The Authors report non conflict of interest.

## Notes

### Competing Interest Statement

The authors have declared no competing interest.

### Author Declarations

All participants provided written informed consent according to the Declaration of Helsinki prior to the study inclusion. This study was approved by the Fondazione IRCCS Istituto Neurologico C. Besta ethics committee.

